# Pregnant women’s perceptions of an ICT-based reminder and monitoring system for antenatal care services in rural coastal Tanzania: A cross- sectional study

**DOI:** 10.64898/2026.09.08.26362454

**Authors:** Johnson Mshangila, Charles Marwa

## Abstract

**Objective:** This study assessed the level of early antenatal care attendance, identified barriers to attendance, and assessed pregnant women’s perceptions of an ICT-based reminder and monitoring system in rural coastal Tanzania.

**Design:** A Cross-sectional study was conducted using structured interviewer administered questionnaires with data collected through kobotoolbox.

**Setting:** This study was conducted at antenatal care clinics of Bagamoyo and Mkuranga District Hospitals, Coastal Region, Tanzania, from April to May 2026.

**Participants:** 308 pregnant women who attended antenatal care clinics were systematically sampled from ANC registers.

**Results:** 31.2% of pregnant women initiated antenatal care within the first 12 week of gestation. Prolonged waiting time at antenatal care clinics was the most prominent barrier, followed by distance from health facility and feeling healthy. Four constructs of the Health Belief Model were associated with positive perception toward the ICT-based reminder and monitoring system; High perceived susceptibility (aPR = 3.517; 95%CI: 1.846–6.701), high perceived benefit (aPR = 2.697; 95%CI: 1.472–4.945), high self-efficacy (aPR = 1.498; 95%CI: 1.285–1.872), high cue to action (aPR = 1.941; 95%CI: 1.121–3.359).

**Conclusion:** Early antenatal care attendance in rural settings still falls below recommended WHO level, attributed to individual, accessibility, and health system barriers. The ICT-based reminder and monitoring system was perceived favorably by pregnant women and has the potential to improve timely antenatal care attendance and service delivery.

**Key point:** *Questions:* What is the level of early antenatal care attendance among pregnant women in rural coastal Tanzania? What are the barriers faced by pregnant women who attended antenatal care clinics? What are pregnant women’s perceptions of an ICT-based reminder and monitoring system to support their attendance at antenatal care services?

*Finding:* Only 31.2% of 308 pregnant women initiated antenatal care within 12 weeks of gestation. The predominant barriers to antenatal care attendance were prolonged waiting time, distance to facility, and feeling healthy. High perceived susceptibility, perceived benefit, self-efficacy, and cue to action were positively associated with positive perception of the ICT-based reminder and monitoring system.

*Meaning:* Pregnant women in rural coastal Tanzania had positive perceptions toward an ICT-based reminder and monitoring system and viewed it as a potentially useful approach for improving antenatal care uptake when combined with health education and the reduction of health facility structural barriers.

## 1. Introduction

Neonatal mortality remains a public health problem in Tanzania despite on going efforts to improve maternal and child health services [1,2]. Progress in reducing neonatal mortality has been slow, particularly in rural areas where access to quality antenatal care (ANC) services is limited [3,4]. Early ANC attendance enables pregnant women to acess timely delivery of essential interventions, including tetanus immunization, malaria and HIV prevention, nutritional supplementation, birth preparedness, and early detection of pregnancy complications, all of which contribute to improved maternal and neonatal outcomes [5,6].

Despite national policies promoting maternal and child health, including free maternal health services and community health programs, early ANC utilization remains low in many rural districts [7]. Delayed ANC initiation has been attributed to limited awareness, forgetfulness, long travel distances, socio-cultural beliefs and weak follow-up systems, resulting in missed opportunities for preventive care and increased risks of adverse neonatal outcomes [3,4], [6].

Advances in information and communication technology (ICT) have created new opportunities to improve maternal and neonatal health in low-resource settings [8]. ICT-based reminder and monitoring systems, which use mobile phone-based interventions such as SMS reminders and digital tracking tools, are designed to notify pregnant women about scheduled ANC visits, provide health education messages, and support monitoring of ANC attendance, thereby enhancing appointment adherence, facilitating follow-up, and strengthening continuity of care [8]. The rapid expansion of mobile phone ownership in Tanzania, including in rural areas, provides a practical platform for integrating ICT into routine maternal healthcare delivery [9–11].

Evidence on the pregnant women’s perceptions of ICT-based reminder and monitoring systems to support early ANC attendance remains limited, particularly in rural Tanzania. Generating context-specific evidence on pregnant women’s perceptions is essential to understand how such interventions are viewed and to inform their design, implementation, and potential scale-up to strengthen ANC utilization and ultimately contribute to improved maternal and neonatal health outcomes in Tanzania and similar resource-limited settings. Therefore, this study assessed the level of early ANC attendance, identified barriers to ANC attendance, and assessed pregnant women’s perceptions of an ICT-based reminder and monitoring system in rural coastal Tanzania. The findings may inform future development and implementation of digital health interventions to support timely ANC attendance in Tanzania.

## 2 Methods

### 2.1 Study Design

A cross-sectional study employed a approach to assess the level of early ANC attendance, barriers to attendance, and perceptions of an ICT-based reminder and monitoring system among pregnant women who attended ANC clinics at Bagamoyo and Mkuranga District Hospitals from April to May 2026.

### 2.2 Study setting

The study was conducted at Bagamoyo and Mkuranga district hospitals, located in coastal region of Tanzania. These district hospitals were purposively selected because they serve predominantly rural population andhad high numbers of maternal health services users in coastal region [12]. According to the the District Health information system (DHIS 2) reports, both hospitals reported relative low ANC and PNC attendance during 2025[13]. Both hospitals have Reproductive and Child health (RCH) units that provide routine antenatal and postnatal care services and therefore served as the recruitment sites for this study.

### 2.3 Study population

The study population consisted of pregnant women who attended ANC clinics at Bagamoyo and Mkuranga district hospitals.

### 2.4 Sample size

The sample size for the quantitative component was calculated using Cochran’s formula for cross-sectional studies [14], using a 95% confidence level (Z = 1.96), an estimated prevalence of early ANC attendance of 24% from a previous Tanzanian study [15], and a 5% margin of error. The minimum required sample size was 280 pregnant women. After adjusting for a 9% non-response rate based on the pilot study, the final sample size was 308 participants. The sample was proportionally allocated between Mkuranga and Bagamoyo district hospital based on their average monthly ANC attendance obtained from quarterly DHIS2 reports [13].

### 2.5 Sampling procedure

A systematic random sampling technique was used to select the study participants. The ANC register at Bagamoyo and Mkuranga district hospitals served as the sampling frames for eligible pregnant women who attended ANC clinic during the study period. A random starting point was selected from each sampling frame, after which every second eligible pregnant woman was selected until the required sample size for each hospital was reached. This approach helped to minimize the risk of selection bias during participant selection.

### 2.6 Inclusion and Exclusion criteria

The pregnant women who attended ANC clinics at Bagamoyo and Mkuranga District Hospitals during the study period who provided consent. Pregnant women who were critically ill were excluded from the study.

### 2.7 Variables

ANC attendance was measured as a binary variable, categorized as early or late ANC attendance. Based on the ANC guidelines [16], early ANC attendance was defined as initiation of ANC before 12 completed weeks of gestation, while late ANC attendance was defined as initiation of ANC at 12 weeks of gestation or later.

Socio-demographic and obstetric characteristics included age, marital status, education level, occupation, parity, and gravidity. Barriers to early ANC attendance were assessed using items informed by the Three Delays Model, which categorizes barriers into delays in seeking care, delays in reaching health facilities, and delays in receiving adequate care at health facilities.

Pregnant women’s perceptions of the ICT-based reminder and monitoring system were assessed using perception-related items adapted from the Health Belief Model (HBM). The items assessed key HBM constructs, including perceived susceptibility, perceived severity, perceived benefits, perceived barriers, self-efficacy, and cues to action. Responses to the perception items were scored and summed to generate a composite perception score and categorized based on a predefined cut-off point.

### 2.8 Data collection

Data were collected using a structured interviewer administered questionnaire. The questionnaire was digitized using the KoboToolbox data collection platform, which allowed for efficient data entry, real time validation, and secure storage [17].

The data collection tools for this study were pre-tested at Kisarawe district hospital, and the review and inputs from the pretest were used to refine the actual data collection tools to make it more effective. Kisarawe district hospital was selected because the community it serves resemble the similarity to the study site hospitals in terms of the cultural and coastal geographical location.

### 2.9 Data collection procedure

Data collection was carried out by the principal investigator (PI) and three research assistants with nursing backgrounds. The research assistants were trained for one week prior the actual data collection activity on the study objectives, ethical consideration, proper use of data collection tools and digital data entry procedures.

Data collected via KoboToolbox were submitted electronically to a password protected KoboToolbox account accessible only to the PI. Appropriate measures were taken to maintain data privacy, confidentiality and secure storage.

The reliability of the study questionnaire was assessed using Cronbach’s alpha. The overall Cronbach’s alpha coefficient was 0.839 indicating good internal consistency.

### 2.10 Ethical approval

Ethical approval was obtained from the College of Business Education Institutional Review Board (Ref. No. 04.8291.02.01.2024), and administrative permissions were secured from the respective hospitals. The study was conducted in accordance with the Declaration of Helsinki. All eligible participants were informed about the study objectives, procedures, potential risks, and benefits, and were assured of their right to refuse or withdraw participation at any time without consequences. Written informed consent was obtained from participants before data collection, with verbal consent applied where applicable.

### 2.11 Patient and public involvement

Patients and the public were not involved in the design, conduct, reporting or dissemination plans of this research.

### 2.12 Data management and analysis

Data exported in Microsoft excel format from KoboToolbox and subsequently imported into Stata version 15 software. The dataset was cleaned and checked for the missing data and normal distribution of the variables.

Descriptive analysis was used to analyse the socio-demographic characteristics of the study participants, level of ANC attendance among pregnant women and barriers to it and findings were presented into percentages.

Bivariate analysis using Generalized Linear Models (GLM) was performed to identify factors associated with pregnant women’s perceptions of the ICT-based reminder and monitoring system.Variables with a p-value less than 0.2 in the bivariable analysis were included in the multivariable GLM for binomial family with log link to control for potential confounders. Statistical significance was set at p-values less than 0.05.

## 3 Results

### 3.1 General description of study respondents

A total of 308 pregnant women attended ANC services at Mkuranga and Bagamoyo district hospitals were enrolled in this study. The participant’s ages ranged from 16 to 43 years, with a mean age of 27 years (SD ± 5.65). The majority of participants fell within the 25–34 years (50.97%). Regarding marital status, most participants were married (73.05%). In terms of education attainment, nearly half had attained secondary school education (48.05%) and occupation status nearly half of the participants were self-employed (49.68%).

On the obstetric characteristics, more than half of the participants were multigravida (55.84%), indicating previous experience of two or more pregnancies, while the majority were nulliparous indicating they had not previously delivered a child (50.65%). Most participants were in the third trimester of pregnancy (52.60%), and the majority had attended fewer than four ANC visits (69.48%). Slightly more than half of the respondents reported living less than 5 km from the district hospital (51.95%). The socio-demographic characteristics of the pregnant women are summarized in Table 1.

**Table 1:**
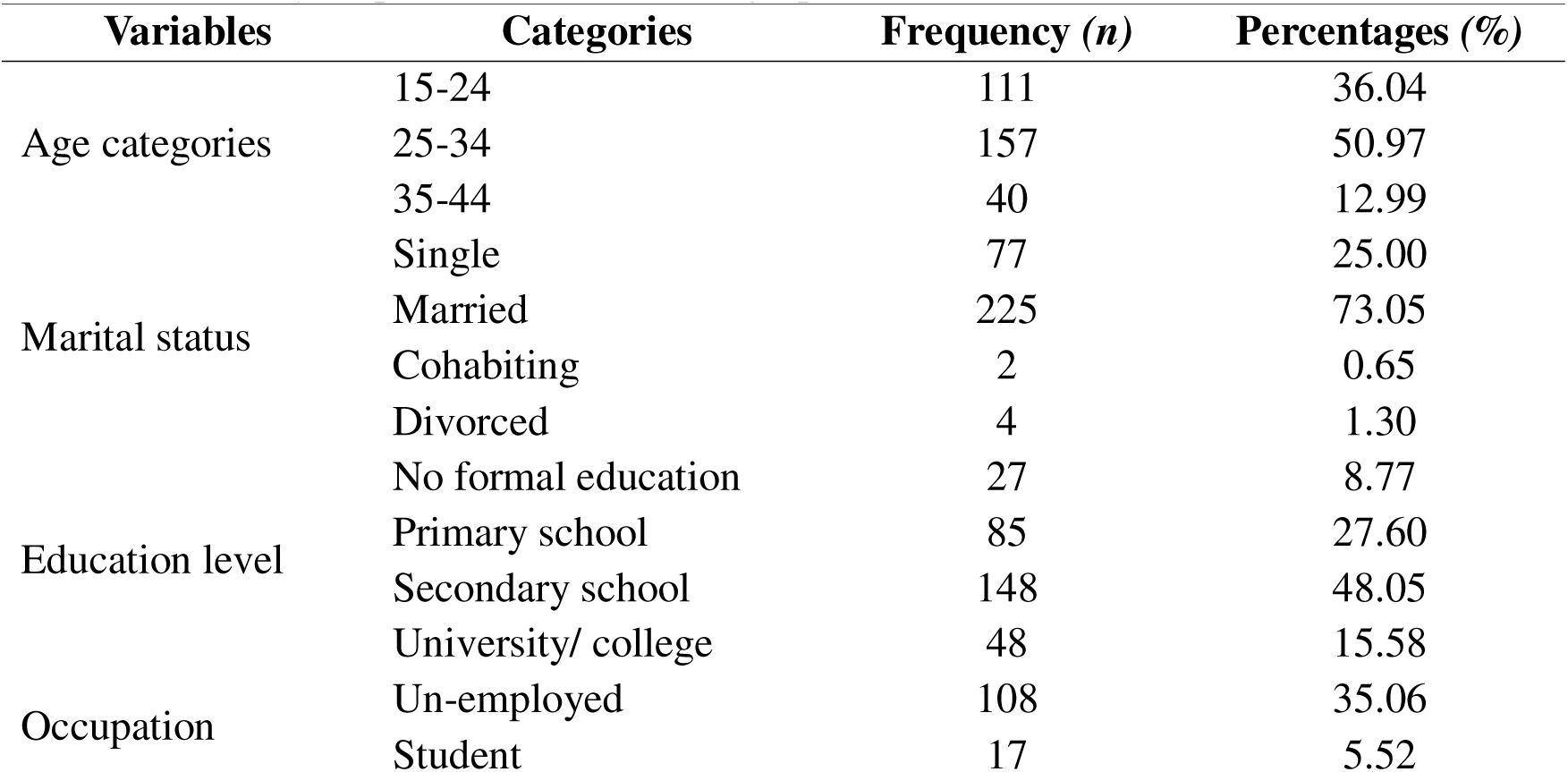
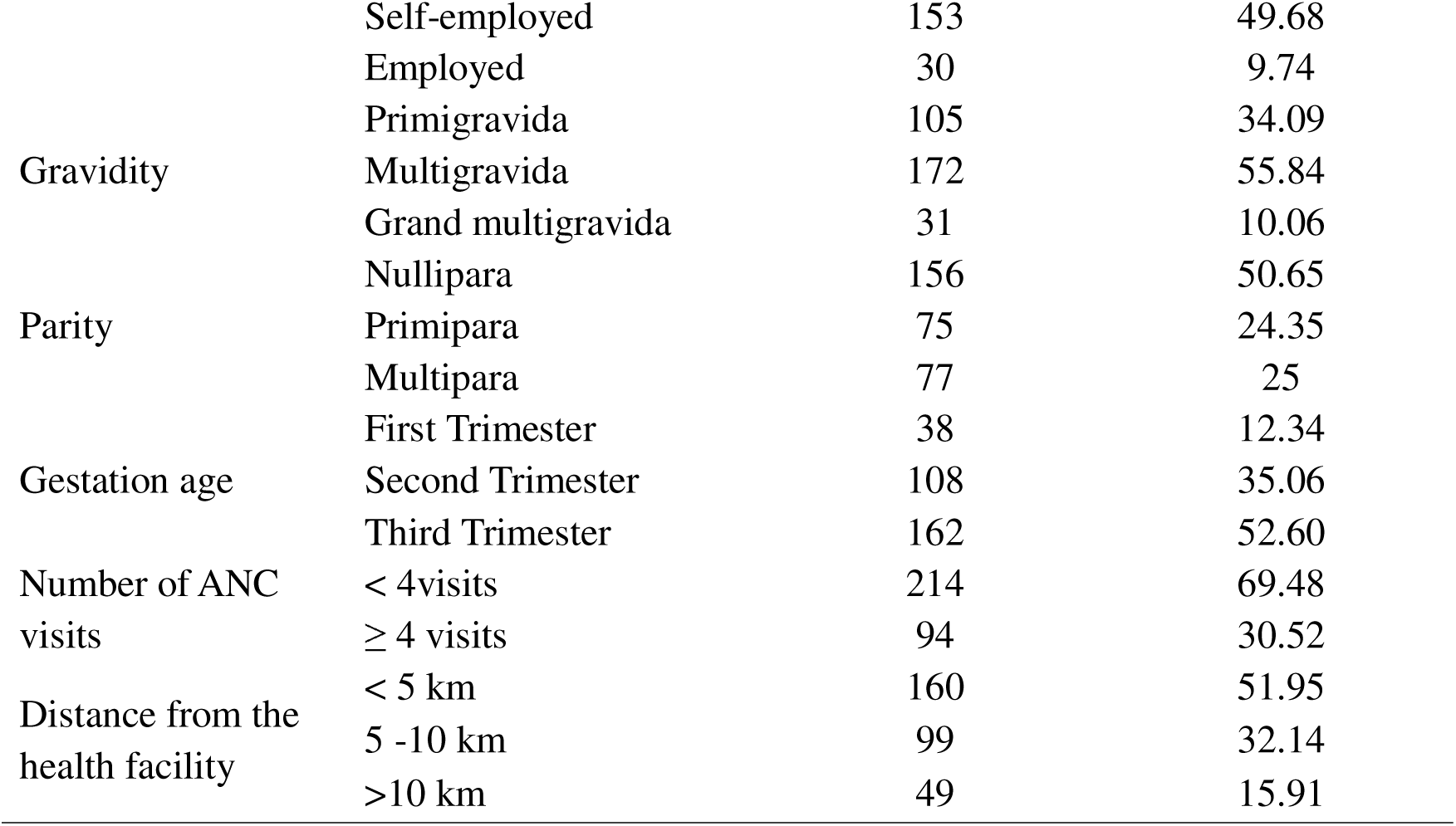
Study respondent’s socio-demographic characteristics.

#### Level of antenatal care attendance among pregnant women

The study findings revealed that only 31.2% of pregnant women initiated antenatal care early within the first trimester, while a substantial proportion of pregnant women initiated ANC late, beyond the World Health Organization (WHO) recommended period of the first 12 weeks of gestation. Figure 2 describes the distribution of pregnant women according to the timing of ANC initiation.

#### Barriers to antenatal care attendance among pregnant women

Barriers to ANC attendance among pregnant women were assessed using the three delays model. The three delays model is a widely recognized framework comprising 3 domains which are delay in decision to seek care, delay in reaching health facilities, and delay in receiving adequate healthcare. The mean scores of identified barriers ranged from 2.18 to 3.18, reflecting varying levels of barriers across the 3 domains that impeded timely ANC attendance among pregnant women. The mean scores of barriers to ANC attendance among pregnant women is presented in figure 3.

#### Delay in Decision to Seek Care (Individual Factors)

Under this domain, feeling healthy had a relatively high mean score of 2.95, indicating that many pregnant women delayed attending ANC because they perceived themselves as healthy and therefore saw no immediate need for attending the ANC during the first 12 weeks of gestation.

Similarly, less necessity to attend ANC early had a mean score of 2.72, indicating inadequate awareness regarding the importance of early ANC initiation.

The cultural and family beliefs had a lower mean score of 2.53, although it still contributed moderately to delayed ANC attendance. Under this domain the personal perceptions and knowledge gaps were major contributors to delayed decision-making for ANC attendance.

#### Delay in Reaching Health Facility (Accessibility Factors)

Under this domain, the health facility being far from home recorded a mean score of 2.72, making it the major barrier within this domain. This indicates that geographical distance from the district hospital remains a challenge for pregnant women seeking ANC services.

Transport cost and travel time had mean scores of 2.33 and 2.31 respectively, suggesting moderate barriers on ANC attendance. Under this domain of the accessibility factors, the physical accessibility and transportation challenges hinder timely utilization of ANC services among pregnant women.

#### Delay in Receiving Adequate Healthcare (Health System Factors)

Under this domain the waiting time had the highest mean score overall (3.18), indicating that prolonged waiting at ANC clinic was the most prominent barrier affecting ANC attendance among pregnant women.

Not being satisfied with the quality of services at the ANC clinic had a mean score of 2.31, while healthcare workers not being supportive had the lowest mean score of 2.18.

Under this domain of health system factors, the waiting time and the satisfaction challenges hinder timely utilization of ANC services among pregnant women.

#### Factors associated with perceptions of the ICT-based reminder and monitoring system

Several factors were associated with perceptions of the ICT-based reminder and monitoring system for antenatal care services as summarized in Table 2. Overall, five out of six Health Belief Model constructs were found to be significantly associated with pregnant women’s perceptions of the ICT-based reminder and monitoring system.

**Table 2:** Factors associated with perception of an ICT-based ANC reminder and monitoring system in bivariate and multivariable analysis.

| Variable | Category | Bivariable analysis |  |  | Multivariable analysis |  |  |
| --- | --- | --- | --- | --- | --- | --- | --- |
|  |  | cPR | 95% CI | P-Value | aPR | 95% CI | P-Value |
| Perceived susceptibility | Low | Ref |  |  | Ref |  |  |
|  | High | 1.274 | 1.099-1.477 | 0.001 | 3.517 | 1.846-6.701 | **0.001\*** |
| Perceived benefit | Low | Ref |  |  | Ref |  |  |
|  | High | 1.227 | 1.055-1.426 | 0.008 | 2.697 | 1.472-4.945 | **0.001\*** |
| Perceived severity | Low | Ref |  |  | Ref |  |  |
|  | High | 0.866 | 0.744-1.009 | 0.066 | 0.566 | 0.312-1.025 | 0.060 |
| Perceived barrier | Low | Ref |  |  | Ref |  |  |
|  | High | 0.860 | 0.7388-1.002 | 0.053 | 0.318 | 0.159-0.635 | **0.001\*** |
| Cue to action | Low | Ref |  |  | Ref |  |  |
|  | High | 1.204 | 1.039-1.396 | 0.014 | 1.941 | 1.121-3.359 | **0.018\*** |
| Self-efficacy | Low | Ref |  |  | Ref |  |  |
|  | High | 0.806 | 0.695-0.934 | 0.004 | 1.498 | 1.285-1.872 | **0.015\*** |

**Note:** CI=Confidence Interval; cPR=Crude Prevalence Ratio; aPR=Adjusted Prevalence Ratio \*significant p<0.05.

Perceived susceptibility to pregnancy related complication was significantly associated with a positive perception toward the ICT based reminder and monitoring system. Pregnant women with high perceived susceptibility were 3.517 times more likely to hold a positive perception of the system compared to those with low perceived susceptibility (aPR = 3.517; 95% CI: 1.846–6.701).

Perceived benefit of ANC services was associated with a positive perception toward the ICT based reminder and monitoring system. Pregnant women who perceived high benefit of ANC services were 2.697 times more likely to hold a positive perception of the system compared to those with low perceived benefits (aPR = 2.697; 95% CI: 1.472–4.945).

Perceived barriers to ANC attendance were associated with pregnant women’s perception toward the ICT-based reminder and monitoring system. pregnant women’s who reported high perceived barriers were 0.318 times less likely to have a positive perception of the ICT-based system compared to those who reported low perceived barriers (aPR = 0.318; 95% CI: 0.159–0.635).

Cue to action was associated with women’s perception toward the ICT-based reminder and monitoring system. Pregnant women who reported high cue to action were 1.941 times more likely to have a positive perception of the system compared to those with low cue to action (aPR = 1.941; 95% CI: 1.121–3.359).

Self-efficacy was significantly associated with women’s perception toward the ICT-based reminder and monitoring system. Pregnant women with high self-efficacy were 1.498 times more likely to have a positive perception of the system compared to those with low self-efficacy (aPR = 1.498; 95% CI: 1.285–1.872).

Perceived severity of pregnancy-related complications was not significantly associated with perception toward the ICT-based reminder and monitoring (aPR = 0.566; 95% CI: 0.312–1.025).

## 4 DISCUSSION

This study found that only 31.2% of pregnant women initiated antenatal care within the first trimester, while the majority attended late, falling short of the WHO recommendation of ANC initiation within the first 12 weeks of gestation. This finding reflects a persistent and concerning gap in timely ANC utilization in rural Tanzania settings.

Studies across Tanzania and other sub-Saharan Africa countries have consistently reported low level of early ANC initiation [18–20]. The Tanzania Demographic and Health Survey (TDHS) has consistently reported that early initiation remains low, with many women making their first visit in the second or even third trimester [18]. This pattern is not unique to Tanzania, comparable trends have been documented across 36 sub-Saharan Africa countries with early ANC initiation rates ranging from 14.5% to 68.6%, suggesting that late ANC initiation is a regional rather than isolated challenge [21]. The low rate of early ANC initiation has important implications for maternal and neonatal health, as delayed attendance limits timely access to essential interventions, including complication screening, nutritional supplementation, malaria prevention, and prevention of mother-to-child transmission of infections. This highlights the need for innovative approaches, such as ICT-based reminder and monitoring systems, to promote timely ANC attendance, improve service uptake, and ultimately enhance neonatal outcomes.

This study found that feeling healthy during early pregnancy was the dominant individual-level barrier to timely ANC initiation. Many pregnant women perceived no immediate need to seek care in the absence of symptoms or complications, reflecting a widespread belief that ANC is only necessary when something feels wrong. Studies conducted in Tanzania have reported that women who felt healthy during early pregnancy were significantly less likely to initiate ANC in the first trimester, viewing the visit as unnecessary when no discomfort was experienced [19,20,22]. Similar findings have been reported across sub-Saharan Africa countries which revealed that self-perceived good health as one of the most frequently cited reasons for delayed ANC attendance among pregnant women [23,24]. The consistency of this finding across African settings suggests that misconceptions about ANC being necessary only for sick women are widespread and not limited to a specific context. Addressing this perception requires health promotion strategies beyond encouraging attendance, including community-based education, peer counseling, and engagement of community health workers to emphasize the routine and preventive benefits of ANC and promote early care-seeking behavior.

Distance from health facilities emerged as the most prominent accessibility barrier to ANC attendance in this study. This finding mirrors evidence from across Tanzania and sub-Saharan Africa, where physical distance to health facilities has been consistently and repeatedly identified as one of the most critical structural barriers to timely ANC utilization [25,26]. Studies conducted in rural and urban Tanzania have revealed that women living far from health facilities were significantly less likely to attend ANC early, with distance acting as both a physical and psychological deterrent to care-seeking [19,22,27]. Comparable findings have been reported in East Africa Countries studies, including studies from rural Uganda and Rwanda where geographical remoteness was strongly associated with late or incomplete ANC attendance [28–31]. This finding suggests that facility-based ANC alone may not adequately reach all pregnant women, particularly those in rural and hard-to-reach areas. Expanding community health worker outreach, mobile ANC clinics, and integration of ANC services into community-level primary healthcare could help reduce geographical barriers and improve equitable access to timely ANC.

Prolonged waiting time at health facilities was not only the dominant health system-level barrier but also the single most prominent barrier across all three domains assessed in this study. This finding reflects a significant health system challenge that directly discourages pregnant women from seeking timely and consistent ANC. This is consistent with findings from Tanzania, where studies have reported that long waiting times at reproductive and child health clinics were among the most frequently cited reasons for women either delaying ANC initiation or failing to complete the recommended number of visits [19,22,27]. Studies across Sub-Saharan Africa have consistently identified prolonged waiting time as a key driver of dissatisfaction with ANC services and a barrier to continued utilization [21,32,33]. This finding strongly corroborates the broader body of literature indicating that health system inefficiencies particularly at the point of service delivery remain a critical and under addressed obstacle to maternal health service utilization across Africa. This finding indicates that improving ANC quality is as important as improving physical access. Health system interventions, including appointment scheduling, task shifting, improved clinic organization, and adequate staffing, are needed to reduce waiting times, enhance patient experience, and encourage early and consistent ANC attendance.

This study found that perceived susceptibility was the strongest factor associated with a positive perception toward the ICT-based reminder and monitoring system, with women who felt personally at risk of pregnancy complications being more likely to view the system favorably. This finding is consistent with the studies from Tanzania and other Sub-Saharan Africa countries which revealed that pregnant women who perceived themselves at higher risk of pregnancy complications were significantly more likely to accept and engage with mHealth reminder interventions designed to support ANC attendance [34–36]. This finding implies that health education on the risks of unmonitored pregnancies and delayed ANC attendance may enhance women’s motivation to adopt ICT-based ANC support tools, positioning risk communication as an important strategy for promoting system uptake.

This study found that women who perceived high benefits of ANC services were significantly more likely to have a positive perception toward the ICT-based reminder and monitoring system. This suggests that women who already understand and appreciate the value of ANC for their own health and that of their unborn child naturally extend that positive attitude to tools designed to support timely ANC utilization. Studies conducted in Tanzania examining acceptance of mobile health interventions among pregnant women have consistently revealed that perceived usefulness and benefit of the intervention were strong associated with a positive attitudes toward digital Health tools [35,36]. Similarly, a controlled pilot trial in South Africa demonstrated that pregnant women’s recognition of the direct health benefits of ANC services positively influenced their receptiveness to digital reminder and monitoring systems intended to enhance access to such services [34]. This finding implies that health promotion efforts should extend beyond promoting the ICT system alone and focus on increasing awareness of the benefits of ANC attendance, which may enhance acceptance of supportive digital health technologies.

Pregnant women who reported high perceived barriers to accessing ANC were significantly less likely to have a positive perception toward the ICT-based reminder and monitoring system. This finding suggests that women who face persistent obstacles such as long distances, transportation difficulties, and financial constraints may view a reminder system as insufficient to address the practical challenges that prevent them from attending ANC in the first place. Studies across rural setting in Tanzania and other Sub-Saharan Africa countries have consistently reported that women who experienced significant structural barriers to healthcare access were less likely to perceive mHealth interventions as useful, arguing that digital tools could not substitute for the resolution of underlying access problems [34–36]. This finding implies that ICT-based reminder and monitoring systems alone may not overcome existing barriers to ANC utilization and should be complemented by structural interventions, including transportation support, community health worker outreach, decentralized ANC services, and targeted assistance for vulnerable women.

Pregnant women with a high cue to action were more likely to have a positive perception of the ICT-based reminder and monitoring system. This can be due to the reason that women who are already sensitive and responsive to health-related prompts and motivational triggers in general would naturally be more open to the idea of a system designed to remind and prompt them to attend their ANC appointments. This finding is consistent with studies from across Tanzania which revealed that exposure to health reminders can significantly increase positive attitudes toward digital health interventions in antenatal care utilization [35,36]. Similarly, studies across Sub-Saharan countries have reported that women who received health education and reminders digital system were more likely to initiate ANC early and engage positively with digital health tools, suggesting that cue to action is a consistent facilitator of mHealth acceptance across diverse African contexts [37–39]. This finding implies that integrating health education and reminder strategies with ICT-based systems could strengthen cues to action, thereby improving ANC uptake and enhancing pregnant women’s acceptance of the intervention.

Pregnant women who felt confident in their ability to seek and use ANC services (high self-efficacy) were more likely to view the proposed ICT-based reminder and monitoring system positively, seeing it as a helpful and complementary tool. This can be explained as a confident pregnant woman who already believes she can manage her own health is more open to tools that support and reinforce that behavior. Studies from Tanzania and other Sub-saharan countries similarly found that pregnant women with greater confidence in health-seeking were more likely to attend ANC early and respond positively to health interventions [35,39–41]. The similarity of these findings confirms that self-efficacy consistently influences both ANC utilization and acceptance of digital health tools. This implies that interventions aimed at strengthening women’s confidence through counseling, peer support, empowerment initiatives, and community education may enhance acceptance and uptake of ICT-based health interventions.

### 4.1 Study limitation

Since participants were recruited from health facilities, women who had never attended ANC were not represented in the study; therefore, perceptions and barriers among women who are completely disconnected from ANC services may have been underestimated. Additionally, the use of self-reported data may have introduced recall bias. To minimize this limitation, the study employed clear and concise questionnaires that were developed and pre-tested prior to data collection. Participants were also given sufficient time to seek clarification during data collection to enhance the accuracy and completeness of responses.

## 5 Conclusion

This study found that only 31.2% of pregnant women in rural coastal region of Tanzania initiated ANC within the first trimester, indicating persistently low early ANC utilization below WHO recommendation. Major barriers included perceiving oneself as healthy (individual), distance to facilities (accessibility), and prolonged waiting times (health system).

The ICT-based reminder and monitoring system was generally perceived favorably by pregnant women with positive perception driven by high perceived susceptibility to pregnancy complications, recognition of ANC benefits, responsiveness to health cues, and confidence in health-seeking ability.

The ICT-based reminder and monitoring system presents a promising opportunity to improve timely ANC initiation and maternal and neonatal health outcomes in rural Tanzania, its effectiveness and sustainability depend on complementary investments in community health education, structural barrier reduction, and health system strengthening to drive meaningful and lasting impact.

## Acknowledgements

We acknowledge the pregnant women who accepted to be involved in the study, as well as the hospital administration and the research assistants for their support and cooperation.

## Declarations

### Data availability

Due to institutional restrictions, they are not publicly available but may be accessed upon reasonable request and with permission from the respective hospitals.

### Consent for publication

Not applicable. This manuscript does not contain any individual person’s identifiable data.

## REFERENCE

[1] The United Republic of Tanzania Ministry of Health, “Maternal and Perinatal Death Surveillance and Response (MPDSR) Report for Six Years (2018– 2023),” 2025. Accessed: Aug. 01, 2026. [Online]. Available: https://www.moh.go.tz/storage/app/uploads/public/681/091/752/68109175225d9858947981.pdf

[2] “Health data overview for the United Republic of Tanzania,” 2023. Accessed: Aug. 01, 2026. [Online]. Available: https://data.who.int/countries/834

[3] H. D. Kalter, A. K. Koffi, J. Perin, M. A. Kamwe, and R. E. Black, “Maternal interventions to decrease stillbirths and neonatal mortality in Tanzania: evidence from the 2017-18 cross-sectional Tanzania verbal and social autopsy study,” BMC Pregnancy and Childbirth 2023 23:1, vol. 23, no. 1, pp. 849-, Dec. 2023, doi: 10.1186/S12884-023-06099-Y.

[4] B. A. Ayele, E. Holliday, and C. Chojenta, “Determinants of antenatal care service utilisation in sub-Saharan Africa: an analysis of demographic and health surveys data (2015–2022),” Archives of Public Health, vol. 83, no. 1, p. 189, Dec. 2025, doi: 10.1186/S13690-025-01608-1.

[5] “Child Health and Development.” Accessed: Nov. 18, 2025. [Online]. Available: https://www.who.int/teams/maternal-newborn-child-adolescent-health-and-ageing/child-health/integrated-management-of-childhood-illness

[6] Z. A. Bhutta et al., “Can available interventions end preventable deaths in mothers, newborn babies, and stillbirths, and at what cost?,” The Lancet, vol. 384, no. 9940, pp. 347–370, 2014, doi: 10.1016/S0140-6736(14)60792-3.

[7] A. Baten, R. K. Biswas, E. Kendal, and J. Bhowmik, “Utilization of maternal healthcare services in low- and middle-income countries: a systematic review and meta-analysis,” Syst. Rev., vol. 14, no. 1, p. 88, Dec. 2025, doi: 10.1186/S13643-025-02832-0.

[8] H. Mumtaz et al., “Current challenges and potential solutions to the use of digital health technologies in evidence generation: a narrative review,” Front. Digit. Health, vol. 5, p. 1203945, 2023, doi: 10.3389/FDGTH.2023.1203945.

[9] C. Holst et al., “Development of Digital Health Messages for Rural Populations in Tanzania: Multi- and Interdisciplinary Approach,” JMIR Mhealth Uhealth, vol. 9, no. 9, pp. 213–239, Sep. 2021, doi: 10.2196/25558.

[10] C. Munishi et al., “Acceptability of using mobile Health (mHealth) as an intervention tool for people with drug use disorders in Tanga, Tanzania,” PLOS Digital Health, vol. 2, no. 9, p. e0000257, Sep. 2023, doi: 10.1371/JOURNAL.PDIG.0000257.

[11] N. O. Theonest et al., “Status and future prospects for mobile phone-enabled diagnostics in Tanzania,” PLOS Digital Health, vol. 3, no. 8, p. e0000565, Aug. 2024, doi: 10.1371/JOURNAL.PDIG.0000565.

[12] The United Republic of Tanzania Ministry of Health, “Annual Health Statistical Tables and Figures 2024,” 2025. Accessed: Aug. 01, 2026. [Online]. Available: https://www.moh.go.tz/storage/app/uploads/public/698/4e8/b7b/6984e8b7b067c606076637.pdf

[13] Ministry of Health - The United Republic of Tanzania, “Data Visualizer | DHIS2.” Accessed: Jun. 09, 2026. [Online]. Available: https://dhis.moh.go.tz/dhis-web-data-visualizer/index.html

[14] J. Beard, “Simple sample size calculations for cross-sectional studies,” South Sudan Medical Journal, vol. 17, no. 4, pp. 213–216, Nov. 2024, doi: 10.4314/SSMJ.V17I4.12.

[15] J. Setonga, L. Chamwali, and E. Mkuna, “Overcoming Barriers, Embracing Opportunities: Antenatal Care Use in Lushoto, Tanzania,” eajahme, vol. 7, no. 1, Jun. 2024, doi: 10.58498/EAJAHME.V7I1.50.

[16] The United Republic of Tanzania Ministry of Health, “Antenatal Care Guidelines,” 2018.

[17] “KoboToolbox.” Accessed: Aug. 01, 2026. [Online]. Available: https://kf.kobotoolbox.org/#/forms/apf4rfYckfo95RjGGAmVy2/data/table

[18] P. Panayides, “Coefficient alpha: Interpret with caution,” *Eur*. J. Psychol., vol. 9, no. 4, pp. 687–696, 2013, doi: 10.5964/EJOP.V9I4.653.

[19] G. R. Mahiti, S. Chombo, and P. Luoga, “Socio-demographic correlates of booking antenatal care in first trimester among pregnant women in Tanzania. Insights from Tanzania demographic health survey 2022,” Frontiers in Reproductive Health, vol. 7, p. 1669621, Oct. 2025, doi: 10.3389/FRPH.2025.1669621/TEXT.

[20] W. B. Lyoba, C. J. Mpambije, and J. D. Mwakatoga, “Unlock the drivers of early ANC visits among pregnant women in Kasulu town council, Tanzania: an institutional cross-sectional study,” Reprod. Health, vol. 22, no. 1, p. 187, Dec. 2025, doi: 10.1186/S12978-025-02162-3.

[21] S. O. Maluka, C. Joseph, S. Fitzgerald, R. Salim, and P. Kamuzora, “Why do pregnant women in Iringa region in Tanzania start antenatal care late? A qualitative analysis,” BMC Pregnancy and Childbirth 2020 20:1, vol. 20, no. 1, pp. 126-, Feb. 2020, doi: 10.1186/S12884-020-2823-4.

[22] A. Z. Alem et al., “Timely initiation of antenatal care and its associated factors among pregnant women in sub-Saharan Africa: A multicountry analysis of Demographic and Health Surveys,” PLoS One, vol. 17, no. 1, p. e0262411, Jan. 2022, doi: 10.1371/JOURNAL.PONE.0262411.

[23] S. Mgata and S. O. Maluka, “Factors for late initiation of antenatal care in Dar es Salaam, Tanzania: A qualitative study,” BMC Pregnancy Childbirth, vol. 19, no. 1, p. 415, Nov. 2019, doi: 10.1186/S12884-019-2576-0.

[24] M. A. Lateef, D. Kuupiel, G. G. Mchunu, and J. D. Pillay, “Utilization of Antenatal Care and Skilled Birth Delivery Services in Sub-Saharan Africa: A Systematic Scoping Review,” Int. J. Environ. Res. Public Health, vol. 21, no. 4, Apr. 2024, doi: 10.3390/IJERPH21040440.

[25] M. Yihune Teshale, A. Bante, A. Gedefaw Belete, R. Crutzen, M. Spigt, and S. E. Stutterheim, “Barriers and facilitators to maternal healthcare in East Africa: a systematic review and qualitative synthesis of perspectives from women, their families, healthcare providers, and key stakeholders,” BMC Pregnancy Childbirth, vol. 25, no. 1, Dec. 2025, doi: 10.1186/S12884-025-07225-8.

[26] C. J. Okafor et al., “Barriers to Early Antenatal Care: Insights from Muembeladu Hospital, Zanzibar,” Asian Journal of Pregnancy and Childbirth, vol. 8, no. 1, pp. 386–396, Oct. 2025, doi: 10.9734/AJPCB/2025/V8I1176.

[27] S. A. O. Mohamed et al., “Barriers to Antenatal Care Attendance in Developing Countries: A Systematic Review,” Cureus, vol. 17, no. 10, p. e95342, Oct. 2025, doi: 10.7759/CUREUS.95342.

[28] E. T. Konje, J. Hatfield, R. Sauve, S. Kuhn, M. Magoma, and D. Dewey, “Late initiation and low utilization of postnatal care services among women in the rural setting in Northwest Tanzania: a community-based study using a mixed method approach,” BMC Health Services Research 2021 21:1, vol. 21, no. 1, pp. 635-, Jul. 2021, doi: 10.1186/S12913-021-06695-8.

[29] C. S. Uldbjerg, S. Schramm, F. O. Kaducu, E. Ovuga, and M. Sodemann, “Perceived barriers to utilization of antenatal care services in northern Uganda: A qualitative study,” Sexual & Reproductive Healthcare, vol. 23, p. 100464, Mar. 2020, doi: 10.1016/J.SRHC.2019.100464.

[30] H. Nalubwama et al., “Delayed pregnancy disclosure, attributed social factors and implications for antenatal care initiation: a qualitative study among Ugandan women and their partners,” Reproductive Health, vol. 22, no. 1, Dec. 2025, doi: 10.1186/S12978-025-02040-Y.

[31] O. Tengera et al., “Barriers hindering attendance and adherence to antenatal care visits among women in rural areas in Rwanda: An exploratory qualitative study,” PLoS One, vol. 20, no. 5, p. e0323762, May 2025, doi: 10.1371/JOURNAL.PONE.0323762.

[32] K. J. Irene and Dr. R. Okova, “Factors Influencing Late Initiation of First Antenatal Care Services Among Pregnant Women in Kicukiro District, Rwanda 2025,” International Journal of Healthcare Sciences, vol. 13, no. Issue 2, pp. 317–329, Mar. 2026, doi: 10.5281/ZENODO.17661656.

[33] K. U. Mare et al., “Late initiation of antenatal care visit amid implementation of new antenatal care model in Sub-Saharan African countries: A multilevel analysis of multination population survey data,” PLoS One, vol. 20, no. 1, p. e0316671, Jan. 2025, doi: 10.1371/JOURNAL.PONE.0316671.

[34] D. Zenbaba, D. O. Okeke-Obayemi, S. R. Okeke, and S. Yaya, “Why Do First Time Mothers in Sub Saharan Africa Delay Their First Antenatal Care Visit? Evidence From Demographic and Health Surveys (2019–2022) From 18 Countries—A Cross Sectional Study,” Health Sci. Rep., vol. 8, no. 11, p. e71468, Nov. 2025, doi: 10.1002/HSR2.71468.

[35] R. Sewpaul, K. Resnicow, R. Crutzen, N. Dukhi, and P. Reddy, “A Tailored mHealth Intervention for Improving Antenatal Care Seeking and Its Determinants Among Pregnant Adolescent Girls and Young Women in South Africa: Pilot Randomized Controlled Trial,” JMIR Mhealth Uhealth, vol. 13, p. e59144, 2025, doi: 10.2196/59144.

[36] S. L. Mwaseba, E. S. Mwang’onda, and F. A. Ngalawa, “Digital Connectivity and Antenatal Care Utilisation in Tanzania: Exploring the Role of Internet Use,” Rural Planning Journal, vol. 27, no. 2, pp. 1–9, Feb. 2025, doi: 10.59557/RPJ.25.2.2025.218.

[37] S. van Pelt et al., “Pregnant women’s perceptions of antenatal care and utilisation of digital health tools in Magu District, Tanzania: a qualitative study,” *Sex*. Reprod. Health Matters, vol. 31, no. 1, p. 2236782, 2023, doi: 10.1080/26410397.2023.2236782.

[38] M. Kante and M. Målqvist, “Effectiveness of SMS-based interventions in enhancing antenatal care in developing countries: a systematic review,” BMJ Open, vol. 15, no. 2, p. e089671, Feb. 2025, doi: 10.1136/BMJOPEN-2024-089671.

[39] C. Kachimanga et al., “Evaluating the Adoption of mHealth Technologies by Community Health Workers to Improve the Use of Maternal Health Services in Sub-Saharan Africa: Systematic Review,” JMIR Mhealth Uhealth, vol. 12, no. 1, p. e55819, Sep. 2024, doi: 10.2196/55819.

[40] M. Habtu et al., “Pregnant women’s and health workers’ perceptions and experiences on the Rwandan ANC digital module intervention at selected health centres,” PLOS Digital Health, vol. 5, no. 2, p. e0001264, Feb. 2026, doi: 10.1371/JOURNAL.PDIG.0001264.

